# Blood and skeletal muscle ageing determined by epigenetic clocks and their associations with physical activity and functioning

**DOI:** 10.1101/2021.02.12.21251629

**Authors:** Elina Sillanpää, Aino Heikkinen, Anna Kankaanpää, Aini Paavilainen, Urho M. Kujala, Tuija H. Tammelin, Vuokko Kovanen, Sarianna Sipilä, Kirsi H. Pietiläinen, Jaakko Kaprio, Miina Ollikainen, Eija K. Laakkonen

## Abstract

The aim of this study was to investigate the correspondence of different biological ageing estimates (i.e. epigenetic age) in blood and muscle tissue and their associations with physical activity (PA), physical function and body composition.

Two independent cohorts were included, whose age span covered adulthood (23–69 years). Whole blood and m. vastus lateralis samples were collected, and DNA methylation analysed. Four different DNA methylation age (DNAmAge) estimates were calculated using genome-wide methylation data and publicly available online tools. A novel muscle-specific methylation age was estimated using the R-package ‘MEAT’. PA was measured with questionnaires and accelerometers. Several tests were conducted to estimate cardiorespiratory fitness and muscle strength. Body composition was estimated by dual-energy X-ray absorptiometry.

DNAmAge estimates from blood and muscle were highly correlated with chronological age, but different age acceleration estimates were weakly associated with each other. The monozygotic twin within-pair similarity of ageing pace was higher in blood (r=0.617–0.824) than in muscle (r=0.523–0.585). Associations of age acceleration estimates with PA, physical function and body composition were weak in both tissues and mostly explained by smoking and sex. The muscle-specific epigenetic clock MEAT was developed to predict chronological age, which may explain why it did not associate with functional phenotypes. The Horvath’s clock and GrimAge were weakly associated with PA and related phenotypes, suggesting that higher PA would be linked to accelerated biological ageing in muscle. This may, however, be more reflective of the low capacity of epigenetic clock algorithms to measure functional muscle ageing than of actual age acceleration.

Based our results, the investigated epigenetic clocks have rather low value in estimating muscle ageing with respect to the physiological adaptations that typically occur due to ageing or PA. Thus, further development of methods is needed to gain further insight into muscle tissue-specific ageing and the underlying biological pathways.

## Introduction

Human ageing is accompanied by a decline in physical activity (PA), partly resulting from a decline in muscle mass and function commonly referred to as sarcopenia [1]. These age-related changes result in decreased mobility, loss of physical independence and increased morbidity and mortality. The precise underlying molecular mechanisms behind these physiological changes remain poorly characterised. It seems clear that regulation of gene expression impacts the PA-related phenotypes of each tissue, and age-related alterations are involved in the ageing process.

Epigenetic modifications are crucial hallmarks of ageing [2]. As an interface between genome and external factors, epigenetics forms a fundamental link between genotype and environment in ageing. DNA methylation is the most widely studied epigenetic mechanism. It may modify gene function without changing the DNA sequence, in part by reacting to the internal and external environment. Some of the alterations in DNA methylation at specific CpG (a cytosine nucleotide followed by a guanine nucleotide) sites correlate with chronological age. This finding, together with technological innovations in the field of microarrays, development of infrastructures for data sharing and adaptation of statistical modelling methods for biological ageing studies, have offered promising tools for identifying epigenetic markers of biological age [3–6]. The novel epigenetic biomarkers, referred to as epigenetic clocks, are weighted sums of DNA methylation at specific CpG sites that can be used as indicators of biological age. Epigenetic clocks are based on penalty regression models in which chronological age or other ageing phenotypes are predicted using genome-wide DNA methylation data, and optimal CpG sites are selected for the age predictor by the algorithm [3].

First-generation clocks, Horvath’s and Hannum’s, were developed to predict chronological age [3,4]. The epigenetic age estimate, DNA methylation age (DNAmAge), predicts differences in mortality better than chronological age after adjusting for several known risk factors [7]. However, these first clocks were criticised for excluding CpGs whose methylation patterns may reflect deviation of epigenetic age from chronological age [5]. Therefore, the next-generation epigenetic clocks were developed to capture CpGs with their methylation associated with the functional stage and chronological age of an individual. PhenoAge was developed using ‘phenotypic age measure’ instead of chronological age as a reference [5], and GrimAge was built using a combination of DNA methylation-based surrogate biomarkers for mortality, such as methylation-based estimates of health-related plasma proteins and smoking pack-years [6]. These two novel epigenetic age estimates predict mortality risk better than Horvath’s and Hannum’s DNAmAge, while the latter ones perform better in predicting chronological age in large datasets [5,6,8].

The epigenetic clocks are developed using blood-based leukocyte DNA, but they seem to work in several tissues [3,5]. There is some evidence suggesting that different tissues and organs in the same individual may vary in terms of their DNAmAge. The cerebellum shows slower ageing compared to other tissues in supercentenarians [9], while ageing of the liver, rather than blood, skeletal muscle or fat tissue, is accelerated in persons with obesity [10]. These observations lend support to the assumption that tissue-specific analyses might yield further insights into age-related pathologies. Muscle is the key tissue for physical functioning and whole-body metabolism, affecting age-related decline and the onset or severity of several age-related diseases, such as type II diabetes and cardiovascular diseases and even mortality [11–14]. Understanding of DNAmAge in muscle is sparse. However, it could be a potential biomarker of the individual ageing process and may offer insights into the biological mechanisms that regulate muscle ageing.

Potential discrepancies in DNAmAge between tissues could also offer explanations for the relevance of different environmental exposures and their influence on health and morbidity. PA is a modifiable behaviour that has the potential to slow down the rate of cellular and molecular damage accumulation [15] and blunt the decline in physiological function with increasing age. The causal molecular and genetic links between PA and reduced age-related disease risk remain elusive, but it is clear that the effects of PA depend on its mode, intensity, duration and frequency and are tissue specific [16,17].

Very recently, the first epigenetic clock for muscle tissue, ‘MEAT’ (Muscle Epigenetic Age Test), was developed to predict ageing in muscle [18]. It has been suggested that this first muscle-specific epigenetic clock can predict chronological age with better accuracy than, for example, Horvath’s DNAmAge clock, which has been previously shown to have reasonable accuracy in many tissues [3]. To the best of our knowledge, MEAT has not yet been investigated in relation to PA or physical function.

The aim of this study was to compare the biological ageing of blood and muscle tissue determined by several epigenetic clocks in two independent cohorts. In addition, we investigated whether ageing in these two tissues is associated with PA and physical function-related phenotypes.

## Results

### Participant characteristics

The characteristics of the study participants, consisting of groups of younger [mean age 32.9 (SD 5.3) years] adults (Finnish twin cohort [FTC] twins), middle-aged [mean 51.8 (SD 2.0) years)] adults (Estrogenic Regulation of Muscle Apoptosis study [ERMA]) and older [mean age 63.7 (SD 4.2) years] adults (FTC twins) are presented in **Table 1**. The ERMA cohort were all women, while 43% of the younger twins and 61% of the older twins were women. The younger and middle-aged adults were better educated, as 40–48% of them had a tertiary-level degree, while in older adults 20% had a tertiary-level degree. In all groups, alcohol use was rather low, and most were light drinkers. A quarter of the younger and 13% of the older twins were current smokers, while only 6% of the middle-aged adults were current smokers, which is due to the fact that smoking is more common among men.

**Table 1.** Descriptive characteristics of the participant groups in two independent cohorts.

|  | **Young adults**<br>FTC, N=93<br>Mean (SD) | **Middle-aged**<br>ERMA, N=47<br>Mean (SD) | **Older adults**<br>FTC, N=46<br>Mean (SD) |
| --- | --- | --- | --- |
| **Socio-economic and life style factors** |  |  |  |
| Age, year | 32.9 (5.3) | 51.8 (2.0) | 63.7 (4.2) |
| Age, range | 23-42 | 48-55 | 57-69 |
| Sex, male/female, n | 62/47 | -/47 | 18/28 |
| Education, n (%)<sup>+</sup> |  |  |  |
| primary | 1 (1.0) | 2 (4.3) | 9 (20.5) |
| secondary | 52 (51.5) | 26 (55.3) | 26 (59.1) |
| tertiary | 48 (47.5) | 19 (40.4) | 9 (20.5) |
| Smoking, n (%) |  |  |  |
| never | 43 (46.2) | 31 (66.0) | 26 (56.5) |
| former | 27 (29.1) | 13 (27.7) | 14 (30.4) |
| current | 23 (24.7) | 3 (6.4) | 6 (13.0) |
| Alcohol, drinks per week<sup>¤</sup> | 4.4 (7.4) | 4.2 (4.5) | 3.1 (5.5) |
| **Body composition<sup>#</sup>** |  |  |  |
| Body mass index, kg/m<sup>2</sup> | 28.7 (5.8) | 26.0 (3.6) | 28.9 (5.7) |
| Fat mass, kg | 30.4 (13.7) | 26.0 (8.2) | 30.9 (9.9) |
| Lean body mass, kg | 51.4 (10.8) | 42.6 (4.5) | 46.1 (9.1) |
| Percentage of fat, % | 34.9 (10.1) | 35.8 (7.0) | 38.5 (8.1) |
| **Physical activity** |  |  |  |
| Sport index | 2.8 (0.99) |  | 2.1 (1.1) |
| Leisure index | 2.9 (0.60) |  | 2.7 (0.80) |
| Work index | 2.8 (0.82) |  | 3.0 (0.51) |
| **Self-reported physical activity, n (%)** |  |  |  |
| Low |  | 6 (12.8) |  |
| Medium |  | 13 (27.7) |  |
| High |  | 28 (59.6) |  |
| **Monitored physical activity** |  |  |  |
| MVPA, min |  | 56.1 (32.2) |  |
| Step counts per day |  | 9 337 (3 378) |  |
| **Physical function phenotypes** |  |  |  |
| VO<sub>2</sub>max, ml/kg/min<sup>*</sup> | 35.2 (8.5) |  |  |
| Maximal workload, W | 195.8 (54.7) |  |  |
| Walking test, 6 min distance (m) |  | 670 (65) |  |
| Hand grip strength (N) |  | 331 (56) |  |
| Knee extension torque (Nm) |  | 486 (95) |  |
| Vertical jumping height (m) |  | 0.19 (0.043) |  |

<sup>+</sup>missing data in education variable N=10, <sup>¤</sup>one drink =12g/ 100% alcohol; <sup>#</sup> Body composition is estimated by dual-energy X-ray absorptiometry; <sup>\*</sup> N=60; MVPA, moderate to vigorous physical activity; VO<sub>2</sub>max, maximal oxygen uptake

All three groups were overweight on average (Table 1). The percentage of fat increased with age, being highest among the older FTC participants. The ERMA participants had the lowest amount of lean mass, which is logical, as this cohort included only women, who tend to be smaller than men.

Regarding PA, the sport and leisure indexes were lower and the work index was higher in older FTC participants than in younger ones. On average the ERMA participants had daily 56 min of moderate-to-vigorous PA (MVPA) and 9400 steps per day, indicating that they were quite active. Maximal oxygen uptake was 35.2 ml/kg/min in younger FTC adults. The walking test, grip and knee extension strength and vertical jumping height were only measured in the ERMA (mean values are presented in Table 1).

### Accuracy of different epigenetic clocks in estimating chronological age

**In blood**, all four clocks showed high correlations between the DNAmAge estimates and chronological age (r=0.904–0.954, all p<0.001, **Figure 1**). The mean deviation between chronological age and predicted age was highest for Hannum’s DNAmAge (7.6 years) and PhenoAge (9.3 years). Lower deviations were observed for Horvath’s DNAmAge and GrimAge (5.6 and 3.6 years, respectively). Horvath’s clock and GrimAge tended to give lower age estimates to older participants than to younger ones, whereas the Hannum’s clock and PhenoAge DNAmAge estimates were consistently lower than the actual chronological age across all age groups (**Figure 1**).

**Figure 1.**
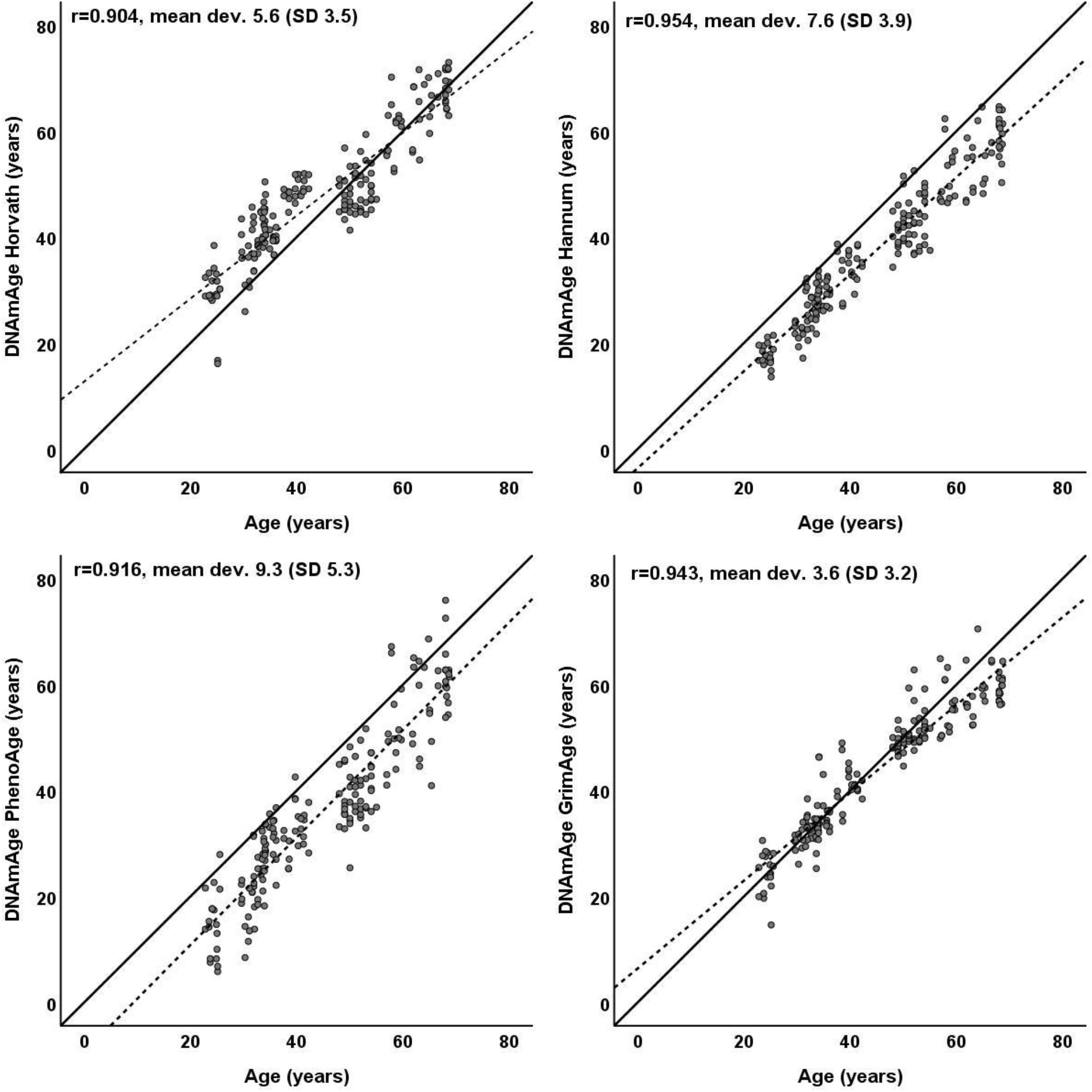
Scatter plots, regression lines (dotted lines), reference lines (solid lines) and Pearson’s correlation coefficients for each of the four predicted DNA methylation ages (DNAmAge) and chronological age in blood. Each dot represents one participant.

**In muscle**, DNAmAge estimated with MEAT showed a higher correlation with chronological age (r=0.962) than DNAmAge estimated with Horvath’s pan tissue clock (r=0.696, **Figure 2**). However, GrimAge, although not developed for muscle tissue, gave age estimates that had an even slightly higher correlation with chronological age (r=0.979). MEAT age estimates showed the highest accuracy (mean deviation 3.2 years). GrimAge age estimates differed from chronological age by 5.0 years, and Horvath’s age estimates differed by 9.5 years. In younger individuals, all clocks tended to give higher muscle age estimates compared with chronological age (**Figure 2**).

**Figure 2.**
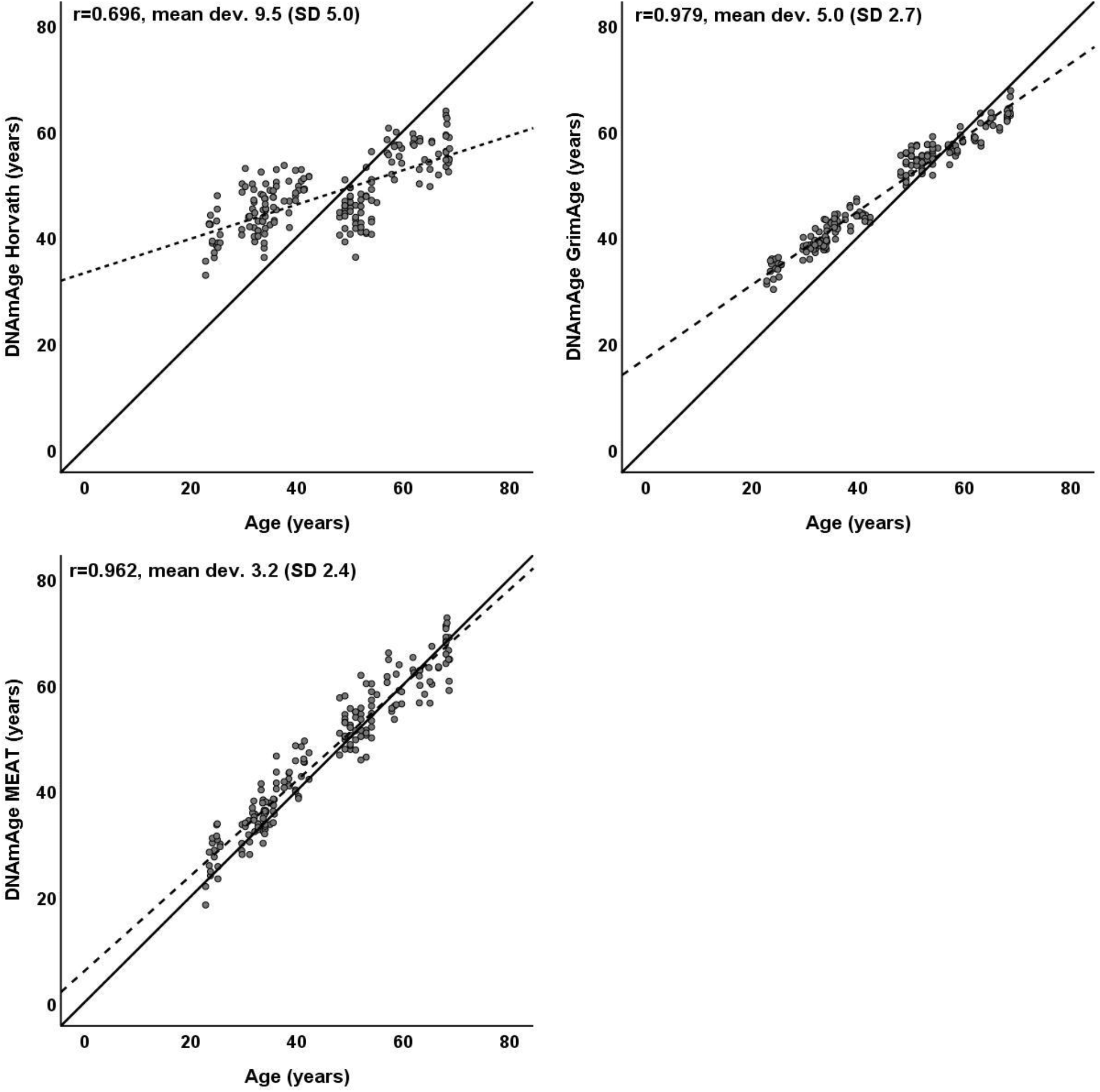
Scatter plots, regression lines (dotted lines), reference lines (solid lines) and Pearson’s correlation co-efficients for each of the three predicted DNA methylation ages (DNAmAge) and chronological age in muscle. Each dot represents one participant.

### Clock and tissue comparison in DNAmAge and age acceleration

In blood, the DNAmAge correlation coefficients were statistically significant between all clocks (r=0.862–0.943) (**Table 2**). Blood DNAmAge estimates also correlated with all the estimates in muscle (r=0.624–0.939). Similarly, DNAmAge estimates in muscle were significantly correlated between all clocks; a highest correlation was observed between MEAT and GrimAge (r=0.940), while the correlations between Horvath’s clock and GrimAge and MEAT were lower (r=0.651 and 0.714, respectively).

**Table 2.** Correlation coefficients between chronological age and different DNAmAge estimates in blood and muscle tissue.

|  |  | Blood DNAmAge |  |  |  | Muscle DNAmAge |  |  |
| --- | --- | --- | --- | --- | --- | --- | --- | --- |
|  |  | Horvath | Hannum | PhenoAge | GrimAge | Horvath | GrimAge | MEAT |
| Blood DNAmAge | Horvath | 1 |  |  |  |  |  |  |
|  | Hannum | 0.928 | 1 |  |  |  |  |  |
|  | PhenoAge | 0.922 | 0.943 | 1 |  |  |  |  |
|  | GrimAge | 0.862 | 0.931 | 0.897 | 1 |  |  |  |
| Muscle DNAmAge | Horvath | 0.732 | 0.703 | 0.624 | 0.632 | 1 |  |  |
|  | GrimAge | 0.86 | 0.937 | 0.878 | 0.939 | 0.651 | 1 |  |
|  | MEAT | 0.876 | 0.927 | 0.874 | 0.913 | 0.714 | 0.940 | 1 |

DNAMAge, DNA methylation age in years; MEAT, muscle epigenetic age test.

All DNAmAge acceleration estimates were also correlated in blood, but the associations were weak to modest (r=0.244–0.566, **Table 3**). In muscle, the strongest correlation was found between Horvath’s and MEAT DNAmAge acceleration estimates (r=0.365). Unlike the DNAmAge results, the blood DNAmAge acceleration values were at most very weakly (all r<0.15) correlated with muscle age acceleration estimates.

**Table 3.**
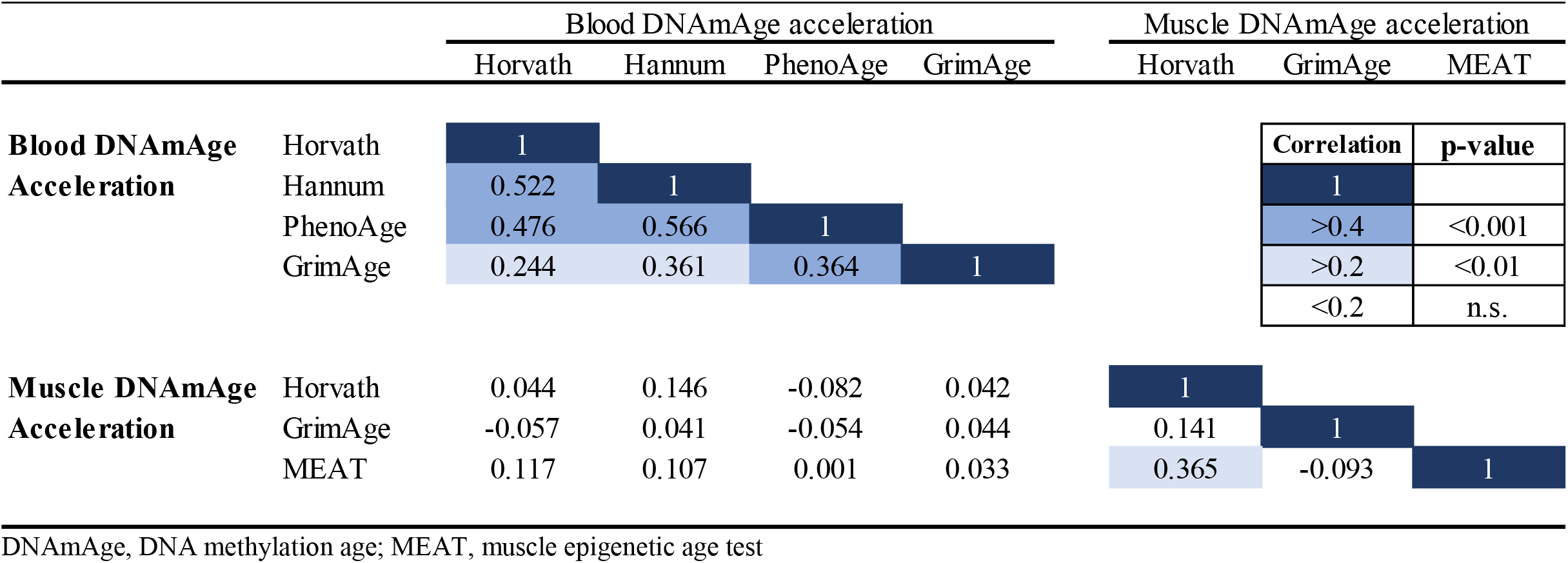
Correlation coefficients between different DNAmAge acceleration estimates in total body (blood) and muscle tissue.

### Within-pair analysis of age acceleration for monozygotic (MZ) pairs

In blood, pairwise correlations were highest in Horvath’s (r=0.807) and Hannum’s age acceleration (r=0.824, **Supplement 1**, Figure 1). Slightly lower within-pair correlations were observed with PhenoAge and GrimAge acceleration (r=0.774 and r=0.617, respectively). In muscle tissue, within-pair analysis showed similar moderate correlations for all three clocks (r=0.523–0.585, **Supplement 1**, Figure 2).

### Associations between epigenetic clocks and physical activity

Associations between DNAmAge acceleration estimates and PA were analysed using both self-reported (FTC and ERMA) and accelerometer-measured activity (ERMA).

In blood, DNAmAge acceleration estimates had minor or non-existent associations with PA (**Supplement 2**, Table 1). In FTC, a weak association of borderline significance was observed between slower age acceleration GrimAge and a higher leisure index. In ERMA, slower age acceleration estimates in GrimAge were evident in the medium-activity group compared to the low-activity participants, and this association attenuated slightly after further adjustments for sex and smoking.

In muscle tissue, the muscle-specific MEAT clock was not associated with PA. Higher age acceleration with Horvath’s clock was only associated with a higher sport index prior to adjustments. Higher DNAmAge acceleration estimated by GrimAge was associated with a higher amount of MVPA and daily steps (**Table 4**), with consistent point estimates (although with larger confidence intervals from the two other clocks).

**Table 4.** Associations between DNAmAge Age Acceleration estimates and physical activity in muscle.

|  |  | Muscle DNAmAge Acceleration |  |  |  |  |  |  |  |  |
| --- | --- | --- | --- | --- | --- | --- | --- | --- | --- | --- |
|  |  | Horvath |  |  | GrimAge |  |  | MEAT |  |  |
| | | $\beta$ (SE) | p-value | R<sup>2</sup> | $\beta$ (SE) | p-value | R<sup>2</sup> | $\beta$ (SE) | p-value | R<sup>2</sup> |
| **FTC& ERMA (N=183)** |  |  |  |  |  |  |  |  |  |  |
| low vs. medium | Model 1 | 0.456 (0.661) | 0.492 | 0.045 | 0.152 (0.273) | 0.578 | 0.022 | 0.120 (0.631) | 0.849 | 0.002 |
| low vs. high |  | **1.842 (0.660)** | **0.006** |  | 0.531 (0.272) | 0.053 |  | 0.338 (0.630) | 0.592 |  |
| low vs. medium | Model 2 | -0.040 (0.667) | 0.951 | 0.111 | 0.059 (0.299) | 0.843 | 0.038 | 0.156 (0.683) | 0.820 | 0.005 |
| low vs. high |  | **1.573 (0.786)** | **0.048** |  | 0.493 (0.268) | 0.068 |  | 0.336 (0.682) | 0.637 |  |
| **FTC (N=136)** |  |  |  |  |  |  |  |  |  |  |
| Leisure index | Model 1 | 0.584 (0.516) | 0.259 | 0.010 | 0.008 (0.189) | 0.967 | 0.000 | -0.763 (0.472) | 0.109 | 0.018 |
|  | Model 2 | 0.483 (0.538) | 0.372 | 0.109 | -0.019 (0.190) | 0.922 | 0.025 | -0.740 (0.523) | 0.161 | 0.020 |
| Sport index | Model 1 | **0.899 (0.338)** | **0.009** | 0.050 | 0.068 (0.123) | 0.592 | 0.002 | -0.221 (0.319) | 0.491 | 0.004 |
|  | Model 2 | 0.756 (0.440) | 0.091 | 0.136 | 0.042 (0.127) | 0.740 | 0.026 | -0.216 (0.377) | 0.569 | 0.006 |
| **ERMA (N=47)** |  |  |  |  |  |  |  |  |  |  |
| **Monitored PA** |  |  |  |  |  |  |  |  |  |  |
| MVPA, min | Model 1 | 0.030 (0.017) | 0.079 | 0.067 | **0.021 (0.008)** | **0.013** | 0.129 | 0.023 (0.016) | 0.163 | 0.043 |
|  | Model 3 | 0.032 (0.018) | 0.075 | 0.080 | **0.022 (0.009)** | **0.014** | 0.161 | 0.021 (0.017) | 0.226 | 0.049 |
| Step counts per day | Model 1 | 0.003 (0.002) | 0.114 | 0.055 | **0.002 (0.001)** | **0.007** | 0.153 | 0.001 (0.002) | 0.548 | 0.008 |
|  | Model 3 | 0.003 (0.002) | 0.105 | 0.068 | **0.002 (0.001)** | **0.006** | 0.189 | 0.001 (0.002) | 0.758 | 0.028 |
| **Self reported PA** |  |  |  |  |  |  |  |  |  |  |
| low vs. medium | Model 1 | 3.22 (1.82) | 0.084 | 0.075 | 0.296 (0.965) | 0.761 | 0.002 | 2.82 (1.79) | 0.122 | 0.056 |
| low vs. high |  | 2.92 (1.66) | 0.085 |  | 0.200 (0.880) | 0.821 |  | 1.58 (1.63) | 0.338 |  |
| low vs. medium | Model 2 | 3.21 (1.87) | 0.093 | 0.082 | 0.209 (0.978) | 0.832 | 0.035 | 2.70 (1.82) | 0.144 | 0.079 |
| low vs. high |  | 2.96 (1.72) | 0.094 |  | 0.101 (0.903) | 0.912 |  | 1.34 (1.68) | 0.428 |  |

Model 1. Linear regression analysis with one predictor. Model 2. As Model one, but adjusted with family relatedness, sex and smoking. Model 3. As model one, but adjusted with smoking. Step counts per day calculated per 100 counts. High activity groups are tertiles formed from moderate to vigorous physical activity (MVPA) and sport indexes. MEAT, muscle epigenetic age test. FTC, Finnish twin cohort; ERMA, Estrogenic Regulation of Muscle Apoptosis study.

In addition to a separate analysis of the two cohorts, we conducted an analysis using both cohorts in the same model, aiming to study associations between DNAmAge estimates and high intensity PA. The ERMA and FTC participants were classified into low-, medium- and high-activity groups based on tertials in the variables MVPA (min) and sport index, respectively. This analysis revealed accelerated ageing among high-activity participants compared to low-activity participants in muscle tissue with Horvath’s clock after adjusting for sex and smoking (**Table 4**).

### Associations between epigenetic clocks and body composition

Associations between blood and muscle DNAm age acceleration estimates and BMI and body composition (fat mass, lean mass and percentage of fat estimated with DXA) were investigated by combining the FTC and ERMA cohorts into the same models.

In blood, the only significant associations were observed between higher age accelerations (Horvath, Hannum and GrimAge) and higher lean mass (**Supplement 1**, Table 2). These associations attenuated substantially after adjusting for sex and smoking.

Associations between muscle DNAmAge acceleration estimates and BMI were not seen. Further, a higher Horvath’s and GrimAge acceleration in muscle was associated in the same direction with lower fat mass and fat percent, even after covariate adjustments (**Table 5**). Higher Horvaths’ age acceleration was only associated with higher lean mass prior to adjusting for sex and smoking. MEAT age acceleration was not associated with body composition.

**Table 5.** Associations between DNAmAge Age Acceleration estimates, body composition and physical function in muscle.

|  |  | Muscle DNAmAge Acceleration |  |  |  |  |  |  |  |  |
| --- | --- | --- | --- | --- | --- | --- | --- | --- | --- | --- |
|  |  | Horvath |  |  | GrimAge |  |  | MEAT |  |  |
| | | $\beta$ (SE) | p-value | R<sup>2</sup> | $\beta$ (SE) | p-value | R<sup>2</sup> | $\beta$ (SE) | p-value | R<sup>2</sup> |
| **FTC& ERMA (N=186)** |  |  |  |  |  |  |  |  |  |  |
| Body mass index, kg/m<sup>2</sup> | Model 1 | -0.049 (0.050) | 0.323 | 0.005 | -0.037 (0.020) | 0.072 | 0.018 | -0.014 (0.047) | 0.759 | 0.001 |
|  | Model 2 | -0.047 (0.048) | 0.334 | 0.014 | -0.036 (0.020) | 0.070 | 0.034 | -0.011 (0.060) | 0.848 | 0.002 |
| Fat mass, kg | Model 1 | **-0.058 (0.023)** | **0.012** | **0.029** | **-0.020 (0.009)** | **0.036** | **0.024** | -0.006 (0.022) | 0.780 | 0.000 |
|  | Model 2 | **-0.050 (0.025)** | **0.048** | **0.009** | **-0.018 (0.008)** | **0.029** | **0.038** | -0.007 (0.025) | 0.788 | 0.003 |
| Lean body mass, kg | Model 1 | **0.084 (0.027)** | **0.002** | **0.045** | 0.012 (0.011) | 0.290 | 0.006 | -0.004 (0.026) | 0.875 | 0.000 |
|  | Model 2 | 0.040 (0.052) | 0.437 | 0.073 | 0.002 (0.019) | 0.909 | 0.018 | -0.018 (0.048) | 0.710 | 0.004 |
| Fat percent, % | Model 1 | **-0.107 (0.029)** | **<0.001** | **0.064** | **-0.030 (0.012)** | **0.014** | **0.032** | 0.008 (0.029) | 0.771 | 0.001 |
|  | Model 2 | **-0.078 (0.038)** | **0.042** | **0.098** | **-0.028 (0.013)** | **0.041** | **0.039** | 0.011 (0.038) | 0.771 | 0.003 |
| **FTC (N=60)** |  |  |  |  |  |  |  |  |  |  |
| Maximal oxygen uptake, ml/kg/min | Model 1 | **0.117 (0.058)** | **0.048** | **0.066** | 0.037 (0.020) | 0.074 | 0.054 | -0.077 (0.044) | 0.082 | 0.051 |
|  | Model 2 | 0.034 (0.069) | 0.626 | 0.195 | 0.028 (0.032) | 0.393 | 0.075 | -0.079 (0.042) | 0.067 | 0.071 |
| **ERMA (N=47)** |  |  |  |  |  |  |  |  |  |  |
| Walking test 6 min, m | Model 1 | 0.002 (0.009) | 0.811 | 0.001 | 0.007 (0.004) | 0.138 | 0.050 | -0.005 (0.008) | 0.506 | 0.009 |
|  | Model 3 | 0.000 (0.009) | 0.959 | 0.010 | 0.006 (0.005) | 0.187 | 0.070 | -0.004 (0.008) | 0.632 | 0.014 |
| Hand grip strength, N | Model 1 | 0.001 (0.100) | 0.904 | 0.000 | 0.000 (0.005) | 0.959 | 0.000 | -0.012 (0.009) | 0.203 | 0.036 |
|  | Model 3 | 0.001 (0.010) | 0.913 | 0.009 | 0.000 (0.005) | 0.973 | 0.034 | -0.012 (0.009) | 0.213 | 0.051 |
| Knee extension torque, Nm | Model 1 | 0.010 (0.006) | 0.098 | 0.066 | 0.004 (0.003) | 0.170 | 0.044 | -0.003 (0.005) | 0.508 | 0.007 |
|  | Model 3 | 0.010 (0.006) | 0.115 | 0.074 | 0.004 (0.003) | 0.285 | 0.066 | -0.004 (0.005) | 0.486 | 0.056 |
| Vertical jumping height, cm | Model 1 | 1.43 (12.95) | 0.915 | 0.000 | 6.246 (6.545) | 0.345 | 0.020 | -11.9 (12.0) | 0.326 | 0.021 |
|  | Model 3 | 0.679 (13.3) | 0.960 | 0.009 | 5.431 (6.664) | 0.420 | 0.048 | -12.8 (12.3) | 0.302 | 0.040 |

Model 1. Linear regression analysis with one predictor. Model 2. As Model one, but adjusted with family relatedness, sex and smoking. Model 3 as model one, but adjusted with smoking. MEAT, muscle epigenetic age test. FTC, Finnish twin cohort; ERMA, Estrogenic Regulation of Muscle Apoptosis study.

### Associations between epigenetic clocks and physical functioning

Associations between blood and muscle DNAmAge acceleration estimates and maximal oxygen uptake were analysed using the younger FTC cohort. No associations were seen in blood, while in muscle tissue higher age acceleration with Horvath’s clock was associated with higher maximal oxygen uptake **(Table 4)**. This was strongly attenuated after adjusting for sex and smoking,

Other analyses of physical function (walking distance, muscle strength and power) were conducted in the ERMA cohort. In blood, higher age acceleration GrimAge was associated with lower knee extension torque before adjusting the model for sex and smoking (**Supplement 1**, Table 2). Other age acceleration estimates in blood and estimates in muscle tissue were not associated with physical function phenotypes.

### Sensitivity analyses

Because our study sample included 50 MZ and 4 DZ twin pairs with discordant BMI, it was important to intervene if the discordance would have affected the observed results. Therefore, we conducted several sensitivity analyses, excluding either the leaner or heavier twin in a pair from the analysis **(Supplement 3)**. In general, the associations were not dependent on whether the analysis was conducted without the heavier or leaner twin in a pair. However, it must be noted that, after excluding the heavier twins, higher MEAT age acceleration was associated with lower maximal oxygen consumption (r=−0.366, p=0.018) compared to a weaker association (r=−0.288) when excluding the leaner twin. In further linear regression analyses with both twins in the model, the association remained significant after adjusting for twin dependency, gender and smoking (β=−0.102, SE 0.046, p=0.034).

## Discussion

Muscle tissue is important with respect to human health, well-being and quality of life. Its proper function is essential for locomotion and other important tasks related to energy metabolism, temperature regulation and recovery from adverse health events. Ageing challenges the optimal functioning of muscle tissue. Typical age-related changes include accelerated decline in muscle function and quantity, which results in problems in functional capacity and health [19]. A method for estimating and predicting muscle ageing would be helpful in identifying persons who are at risk for age-related decline in functional capacity or morbidities, and it could also furnish mechanisms that are potential targets for treatments and interventions to improve muscle functioning among ageing populations. We investigated novel genome-wide epigenetic scores, developed to predict age, age-span and healthy ageing, in blood and muscle tissue [3–6,18]. Our results showed that these clocks are mainly suitable for predicting chronological age from both tissues, but variation in age estimates was not systematically associated with the variation in physical functional phenotypes, which suggests that the clocks lack value for research and health promotion purposes.

### Tissue specificity

We found a strong linear relationship between chronological age and DNA methylation age estimates (i.e. Hannum’s clock, Horvath’s clock, GrimAge, PhenoAge and MEAT) in both blood and skeletal muscle tissues. This was expected, as most of these clocks have been developed to predict chronological age, and age is one component of the GrimAge algorithm. We also replicated the finding regarding the poorer calibration of Horvath’s clock in age estimation in muscle [3,18]. The good overall performance of these clocks in predicting chronological age has been demonstrated repeatedly in previous studies [6,18,20,21]. However, the age acceleration of blood was not associated with the age acceleration in muscle by any of the clocks, suggesting different ticking rates in blood and muscle. We also found age acceleration estimates of blood to be more similar within twin pairs than age acceleration estimates of muscle, suggesting either a larger familial component in blood-based age acceleration estimates or larger heterogeneity in muscle tissue composition. Because we only analysed MZ pairs, we cannot distinguish genetics from environmental effects (e.g. similar occupation or PA) that the twins share in the familial component estimate.

Regarding our hypothesis that PA will slow down age acceleration in muscle, we found that the age acceleration estimates from Horvath’s clock and GrimAge showed a minor but statistically significant positive association with PA measures. These results suggest that higher PA would be linked to accelerated biological ageing in muscle. As PA and ageing typically result in opposite structural and functional changes in muscle, it seems unlikely that PA would accelerate ageing processes in the muscle. Instead, we believe that the findings reflect the low capacity of the epigenetic clock algorithms to measure functional muscle ageing. Possible explanations for this include cell heterogeneity and differences in methylation profiles and structure between white blood cells and muscle tissue. Blood cells represent a constantly dividing cell population, while adult muscle tissue is more heterogeneous. It mainly includes multinucleated post-mitotic myofibers that do not divide, but also a smaller population of quiescent muscle stem cells, called satellite cells, which can re-enter the cell cycle to initiate proliferation and donate nuclei to muscle fibres. Furthermore, a small contribution to the muscle tissue DNA pool is provided by a network of capillaries formed by endothelial and epithelial cell layers as well as a variable, small amount of infiltrated leucocytes and fat and connective tissue cells. PA stimulates the formation of satellite cells, which are essential for muscle regeneration and are responsible for muscle growth [22]. Skeletal muscle cells and satellite cells are differentially methylated, and PA may predispose them to different epigenetic changes depending on age [23]. Satellite cells are important in the regeneration of muscle after cumulative injuries in exercisers, but their functionality declines with increasing age [24]. In addition, ageing and PA typically lead to different structural changes in muscle. In particular, strength training increases the size of muscle cells, while ageing is associated with smaller and weaker muscle cells and changes in the cell-type composition of tissue. However, some structural changes caused by ageing and PA are shared, both resulting in increasing infiltration of fat inside the muscles [25]. However, in the case of PA, infiltrated fat is mainly lipid droplets that do not contain DNA. Muscle tissue samples, although carefully collected and with all visible fat and connective tissue removed, may still contain traces of these other tissues. Whether this explains our finding that both the Horvath’s and GrimAge clock estimates were significantly associated with the amount of body fat in muscle tissue remains unanswered.

### Physical activity

In our study, slower epigenetic age acceleration in blood was weakly or non-significantly associated with higher PA, depending on the epigenetic age estimate and PA measurement method. Substantial associations were observed only with the GrimAge epigenetic clock and with self-reported PA data. The results are in line with those of previous studies, which have reported null findings [21,26] using Horvath’s clock and weak associations using GrimAge [27–29]. The lack of consistent findings regarding the association between epigenetic ageing and PA in our study is likely explained by different features of epigenetic age algorithms and PA data collection methods. We have previously shown that LTPA is associated with slower ageing, while occupational PA is associated with accelerated ageing [27]. This may explain why device-based measures of PA representing whole day PA did not show associations with epigenetic ageing, although they are typically considered more reliable than self-reports in estimating PA.

### Physical functioning

In line with previous studies [30,31], associations between DNAmAge estimates and different physical function phenotypes were non-existent or minor in blood. Within the same individual, different tissues may yield different age estimates [10,32,33]. Some previous studies have suggested that tissue and cell type-specific analyses might be able to track age-related pathologies in target tissues. Muscle is the key tissue affecting age-related decline in physical functioning and PA, and thus we hypothesised that muscle tissue might better reflect biological ageing relevant to functional capacity. We found that the muscle-specific epigenetic clock MEAT is not associated with physical functioning, PA or body composition. MEAT was constructed only to predict chronological age without using any other biological phenotype data and may therefore be independent from molecular alterations caused by changes in the modifiable behavioural phenotypes, such as PA. Muscle-specific clocks need to be further developed taking into account biological and functional parameters, as has been done with respect to the other epigenetic clocks GrimAge and PhenoAge [5,6], which, however, were built for blood tissue only. We also tested the two multi-tissue epigenetic clocks, Horvath’s and GrimAge with muscle tissue. In addition to the previously mentioned association with body fat, the only significant association was between Horvath’s age acceleration and cardiorespiratory fitness. However, this weak association attenuated after adjusting for sex and smoking, suggesting that it originates mostly from biological sex differences and the smoking habits of the participants.

### Strengths and limitations

To the best of our knowledge, this study is the first to examine the associations between epigenetic clocks and PA and physical functioning in muscle tissue, utilising the novel muscle clock MEAT developed by Voisin et al. [18]. Our methods for estimating PA and function covered a wide variety of phenotypes measured in a research laboratory by trained professionals as well as measurement of PA with reliable device-based methods under free-living conditions. By using two independent datasets, we were able to cover a large age span of individuals representing the same genetic ancestry. Since we only included individuals with both blood and muscle data available, we acknowledge that our sample size might not be sufficient to capture the small effect sizes that some phenotypes may have with respect to accelerated ageing. However, the observed trends in the associations between blood GrimAge age acceleration and self-reported PA corresponds to previously reported results in a larger sample [27]. Our sample also included twins with discordant BMI, which may result in larger within-pair differences in methylation profiles. However, we conducted comprehensive sensitivity analyses, excluding either heavier or leaner twins from pairs discordant for BMI from the analysis, and the results were very similar (Supplement 3). Our results should be considered preliminary findings that need to be replicated with other, larger cohorts.

### Conclusions

Both multi-tissue and muscle-specific novel epigenetic clocks seem to perform well in estimating chronological age in muscle tissue. Based on current knowledge and our results, however, these epigenetic clocks have rather low value in estimating muscle ageing with respect to physiological adaptations that typically occur due to ageing or PA. Practical issues related to the sampling of muscle tissue have limited the development of epigenetic research and scoring methods in this area. As age-related functional limitations are expanding rapidly due to increasing life expectancy throughout the world, there is an urgent need to gain more insight into muscle tissue-specific ageing and the underlying biological pathways. This type of knowledge could help the development of medical and lifestyle treatments for age-related functional limitations and muscle pathologies.

## Methods

### Study populations

The data used in the current analysis originate from two Finnish population-based cohort studies: the FTC and the ERMA study.

In brief, **FTC** includes three large cohort studies: 1) study of twins born before 1958, an older twin cohort; 2) in 1975–1979, Finntwin16 (FT16); and 3) in 1983–1987, Finntwin12 (FT12) [34–36]. All three twin cohorts have been studied in an intensive longitudinal manner, and several subprojects have been conducted in laboratory conditions. All twins who had taken part in clinical in-person sub-studies with sampling for both whole blood and muscle tissue, subsequent DNA methylation analyses and phenotype collection were included (N=139) [37,38]. The dataset comprises two age groups: young adults from 23 to 44 years of age (N=93) and older adults from 57 to 69 years of age (N=46). The dataset includes 54 twin pairs, who were discordance for body mass index (BMI; difference more than 3 kg/m^2^) and have participated in the sub-study including laboratory measurements for body composition and physical performance [38]. Zygosity of the twins was confirmed by genetic analysis of multiple polymorphic markers. The majority (45 pairs) were MZ, and only four same sex dizygotic twin pairs were present.

The recruitment process, study design and methodology of the **ERMA** study has been described in detail elsewhere [39]. In brief, the original ERMA cohort includes 47-to 55-year-old women living in the city of Jyväskylä or neighbouring municipalities. The study participants were randomly selected from the national registers of the Digital and Population Data Services Agency, Helsinki (dvv.fi). The ERMA study was designed to reveal how hormonal differences during the menopausal stages affect the physiological and psychological functioning of middle-aged women. The subsample used in the current study (N=47) was randomly selected from participants who had both whole blood and muscle tissue samples available and who did not have missing or low-quality accelerometer data in PA measures.

### Ethics

The FTC data collection and analysis were approved by the ethics committee of the Helsinki University Central Hospital (Dnro 249/E5/01, 270/13/03/01/2008, 154/13/03/00/2011), and the ERMA study was approved by the ethics committee of the Central Finland health care district in 2014 (K-SSHP Dnro 8U/2014). Written informed consent was provided by the participants before the sample collection and PA and functioning measurements.

### Outcome measures

#### Tissue sampling and DNA extraction

Whole blood samples were collected in fasting conditions from the antecubital vein. Muscle biopsy samples were obtained from the mid-part of the m. vastus lateralis, defined as the midpoint between the greater trochanter and the lateral joint line of the knee, by percutaneous needle biopsy under local anaesthesia. Following the removal of all visible fat residues, connective tissue and blood, the biopsy samples were frozen in liquid nitrogen and stored at −150°C until use. Blood white cell and muscle tissue DNA was extracted using a DNeasy Blood & Tissue Kit (Qiagen, Cat No./ID: 69504).

#### Genome-wide methylation analysis

High molecular weight DNA was bisulfite-converted using an EZ-96 DNA Methylation-Gold Kit (Zymo Research, Irvine, CA, USA). Genome-wide DNA methylation was measured using Illumina’s Infinium MethylationEPIC BeadChip (Illumina, San Diego, CA, USA) according to the manufacturer’s instructions. The Illumina BeadChip measures single-CpG resolution DNA methylation levels across the human genome.

#### Data preprocessing

Data were preprocessed using the R package minfi. Detection p-values comparing the total signal for each probe to the background signal level were calculated to evaluate the quality of the samples [40]. Poor-quality samples (mean detection p > 0.01) were excluded from further analysis. Beta values representing CpG methylation levels were calculated as the ratio of methylated intensities (M) to the overall intensities (Beta value=M/(M+U+100), where U is unmethylated probe intensity. Data were normalised using the single-sample Noob normalisation and Beta Mixture Quantile normalisation methods [41,42].

#### Epigenetic clocks

The resulting methylation data were uploaded to an online epigenetic age calculator (https://dnamage.genetics.ucla.edu/new), selecting the option ‘Normalized Data’ to generate the various clock estimates [3]. For blood, the four most commonly used epigenetic age estimates, Horvath’s and Hannum’s DNA methylation age (DNAmAge), PhenoAge and GrimAge, were computed. For muscle tissue, only Horvath’s DNAmAge and GrimAge were used, as Hannum’s clock and PhenoAge were clearly not suitable for muscle tissue according to current knowledge and our preliminary analyses. In addition to these two epigenetic clocks, the novel MEAT was computed using the open access R-package MEAT available on Bioconductor [18]. Samples were removed from all analyses if predicted sex or tissue generated by the algorithm did not match with the actual sex or tissue. Outliers were checked based on cut-off criteria > or < five standard deviations. DNAmAge age acceleration, which describes the rate of individual ageing and is unrelated to chronological age, was calculated for all participants [Age acceleration = residual (linear model (predicted age ~ chronological age)].

#### Physical activity

The Baecke questionnaire was used to assess PA in FTC [43]. The questionnaire consists of 16 items, from which three indexes were calculated: the work index refers to PA at work, the sport index refers to sports participation during leisure time and the leisure-time index refers to PA during leisure time excluding sport activities. All responses were given on a five-point scale, with the exception of questions on main occupation and the types of two main sports. In the original publication, the test–retest reliability of the work index, sport index and leisure-time indices were 0.88, 0.81 and 0.74, respectively [43]. The Baecke questionnaire has been validated for cardiorespiratory fitness among Finnish twins [44].

In the ERMA study, PA was monitored using hip-worn GT3X+ or wGT3X+ ActiGraph accelerometers at 60 Hz (Pensacola, FL) for seven consecutive days [45,46]. The raw acceleration data were converted into 60-second epoch counts and divided into the mean daily duration spent at different PA intensities using tri-axial vector magnitude cut-points of 450 counts per minute (cpm), 2690 cpm and 6166 cpm to separate sedentary from light PA, light from moderate PA and moderate from vigorous PA, respectively; 25000 cpm was used as an upper limit. Mean time spent at MVPA intensity level and daily steps during waking hours were calculated for each participant. Self-reported PA was assessed with a seven-scale questionnaire [47,48]. Due to the low number of participants in some categories, answers were re-categorised as low, medium and high activity.

#### Body composition

Anthropometrics and body composition were measured after overnight fasting. *Body mass index* was calculated as a function of measured body weight and height (kg/m^2^). *Percentage body fat, total body fat mass* and *lean body mass* were measured through dual-energy x-ray absorptiometry (DXA, Lunar Prodigy).

#### Physical functioning

Physical function phenotypes included measures for both cardiorespiratory fitness and upper and lower extremity muscle strength and muscle power. Maximal oxygen consumption was measured only in FTC and other phenotypes and only in the ERMA cohort.

*Maximal oxygen consumption*, which describes the level of cardiorespiratory fitness, was measured with a graded maximal exercise test using an electrically braked bicycle ergometer, as described earlier [49]. The initial workload (40 W for women and 50 W for men) was increased by 40 W (women) or 50 W (men) every 3 min until the maximal exercise capacity was reached (i.e. until volitional exhaustion following generally approved limits) [44]. Oxygen uptake (V? O_2_), carbon dioxide production (VCO2), ventilation, breathing frequency and other standardised respiratory parameters were measured continuously breath by breath with a Vmax spiroergometer (Sensormedics, Yorba Linda, CA). A strict calibration protocol was followed to ensure data quality [44].

*The 6-minute walking test*, in which the participants attempted to walk the longest possible distance in the allotted time, was performed on a 20-m indoor track. Participants were instructed to complete as many laps as possible within 6? min, and the distance they walked was recorded [50]. The 6-min walking test is a performance-based measure of functional exercise capacity, and it can be used as an indicator of exercise tolerance or cardiorespiratory fitness. A standardised protocol and safety guidelines issued by the American Thoracic Society (ATS Committee on Proficiency Standards for Clinical Pulmonary Function Laboratories 2002) were followed.

*Hand grip strength (N)* and *knee extension torque (Nm)* were measured using an isometric dynamometer (Good Strength, Metitur Oy, Jyväskylä, Finland) [51]. To test hand grip strength, the participant’s dominant arm was fixed to the armrest of the dynamometer chair with the elbow flexed at a 90° angle, while knee extension strength was measured at a knee angle of 60° from full extension. During the measurements, participants were encouraged verbally to squeeze the handle or extend their knee to produce maximal contraction. Each participant was allowed to have three to five trials with 1 min rest between trials. The best attempt with the highest 2–3 s lasting peak force value was used in the analysis. Handgrip strength measurement can be used as an indicator of general muscle strength among older people [52]. Maximal Nm was calculated from the best attempt measured in Newtons (N) multiplied by lower leg length (the distance between the lateral joint line of the knee and the midline of a cuff around the ankle) [53].

*Vertical jumping height (cm)* was calculated from flight time during a maximal effort countermovement jump and measured on a contact mat [54]. Three maximal efforts were conducted, and the best result was used in the analysis. Vertical jumping height can be used as an estimate of lower body muscle power (i.e. the ability of the neuromuscular system to produce the greatest possible force as rapidly as possible). Lower extremity muscle power declines earlier and to a greater extent than muscle strength during ageing [55]. It also correlates strongly with mobility among older people [56].

#### Other variables

Sociodemographic variables and life habits were collected with validated questionnaires. These questionnaires have been published previously on the FTC webpage (https://wiki.helsinki.fi/display/twinstudy) and regarding the ERMA study as supplementary digital contents [39]. *Level of education* was classified into primary, secondary and tertiary education groups following the levels of the Finnish education system. *Use of alcohol* was calculated as the number of alcoholic drinks (1 drink is 12 g ethanol) consumed per week, with alcohol consumption computed based on quantity and frequency measures of alcohol beverage use [57,58]. *Smoking* was classified as never, former (abstinence for at least 6 months) and current (daily or occasional) smoker.

#### Statistical analysis

Statistical analyses were carried out with Stata 16 software (StataCorp, Inc. College Station, TX, USA) and R studio (3.6.0). Data are shown as means and standard deviations unless otherwise stated. The mean absolute difference between the predicted DNAmAge and chronological age was calculated to see whether the estimated DNAmAge was younger or older compared to the chronological age. Correlation coefficients were computed to estimate the level of similarity in DNAmAge estimates in different tissues. *DNAmAge age acceleration* was calculated for all participants as the residuals from a linear regression model of the DNAmAge estimate on chronological age. Intraclass correlation coefficients (ICC) were computed for the MZ twin pairs to estimate the level of within-pair similarity. Associations between different DNAmAge acceleration estimates, PA, body composition and PA-related phenotypes were analysed using linear regression analysis, which were further adjusted with sex and smoking. In the models, the within-pair dependency of twin individuals was taken into account using the cluster option in the analysis. The level of significance was set at p≤0.05.

## Data Availability

All twin data used in the current study are located within the Biobank of the National Institute for Health and Welfare, Finland. All the biobanked data are publicly available for use by qualified researchers following a standardized application procedure.

## Acknowledgements

ES was supported by the Academy of Finland (grant number 260001), the Juho Vainio foundation and the Yrjö Jahnsson foundation. VK was supported by the Academy of Finland (275323). KHP was funded by the Academy of Finland (grant numbers 314383 and 266286), the Academy of Finland Center of Excellence in Research on Mitochondria, Metabolism and Disease (FinMIT; grant number 272376), the Finnish Medical Foundation, the Gyllenberg Foundation, the Novo Nordisk Foundation (grant numbers NNF17OC0027232 and NNF10OC1013354), the Finnish Diabetes Research Foundation, the Finnish Foundation for Cardiovascular Research, Government Research Funds, the University of Helsinki and Helsinki University Hospital. JK was supported by the Academy of Finland (213506, 308248, 312073, 336823), EC FP5 GenomEUtwin, NIH NIH/NHLBI (grant HL104125), EC MC ITN Project EPITRAIN and Sigrid Juselius Foundation. MO was supported by the Academy of Finland (297908 and 251316), EC MC ITN Project EPITRAIN, University of Helsinki Research Funds and Sigrid Juselius Foundation. EKL was supported by the Academy of Finland (309504 and 314181) and the Juho Vainio Foundation. The Gerontology Research Center is a joint effort between the University of Jyväskylä and the University of Tampere.

## Author information

ES, EKL and MO designed the study, AH and AK prepossessed methylation data and formed epigenetic age estimates. ES and AH performed the analysis and interpretation of the data and drafted the first version of the manuscript. EKL, JK, UMK, MO, AP and KHP participated in the data interpretation and planning the statistical analyses. VK, ELK, SS, UMK and THT designed and collected the ERMA phenotype dataset. JK, KHP and MO designed and collected the FTC dataset. All authors have been involved in drafting the manuscript or revising it critically for important intellectual content; they have approved the analysis done, as well as have given final approval of the version to be published.

## Availability of data and materials

### Abbreviations

BMI: body mass index, kg/m^2^
CpG: a cytosine nucleotide followed by a guanine nucleotide
DNAmAge: DNA methylation age
ERMA: Estrogenic Regulation of Muscle Apoptosis study
FTC: Finnish twin cohort
ICC: Intraclass correlation coefficients
LTPA: leisure time physical activity
MEAT: muscle epigenetic age test
MVPA: a moderate-to-vigorous physical activity
MZ: monozygotic
PA: physical activity

## Consent for publication

Not applicable.

## Supplementary files

Supplementary file 1. Within-pair correlations in age acceleration in blood and in muscle.

Supplementary file 2. Associations between DNAmAge age acceleration estimates and body composition and physical activity in blood.

Supplementary file 3. Sensitivity analyses related to twin pair discordance in body mass index.

