## Supplemental files for "Blood and skeletal muscle ageing determined by epigenetic clocks and their associations with physical activity and functioning"

**Supplementary file 1. Within pair correlations in age acceleration in blood and in muscle.**

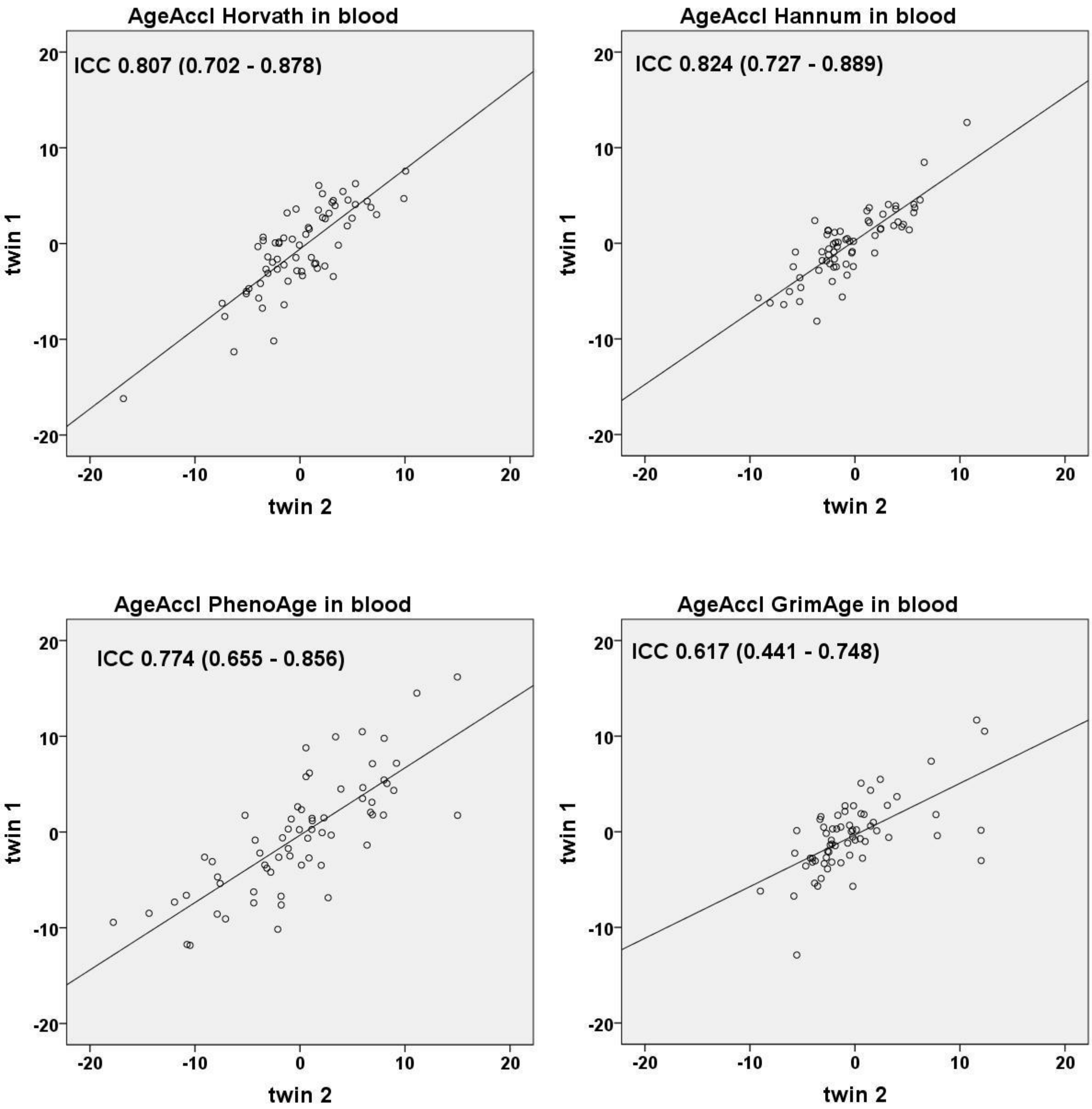


S1_Figure 1. Within-pair (ICC) correlations, their 95% confidence intervals and regression lines of twin1 on twin2 of four age acceleration estimates (AgeAccl) in monozygotic twin pairs (N=65 pairs) in blood. Each dot represents one twin pair.

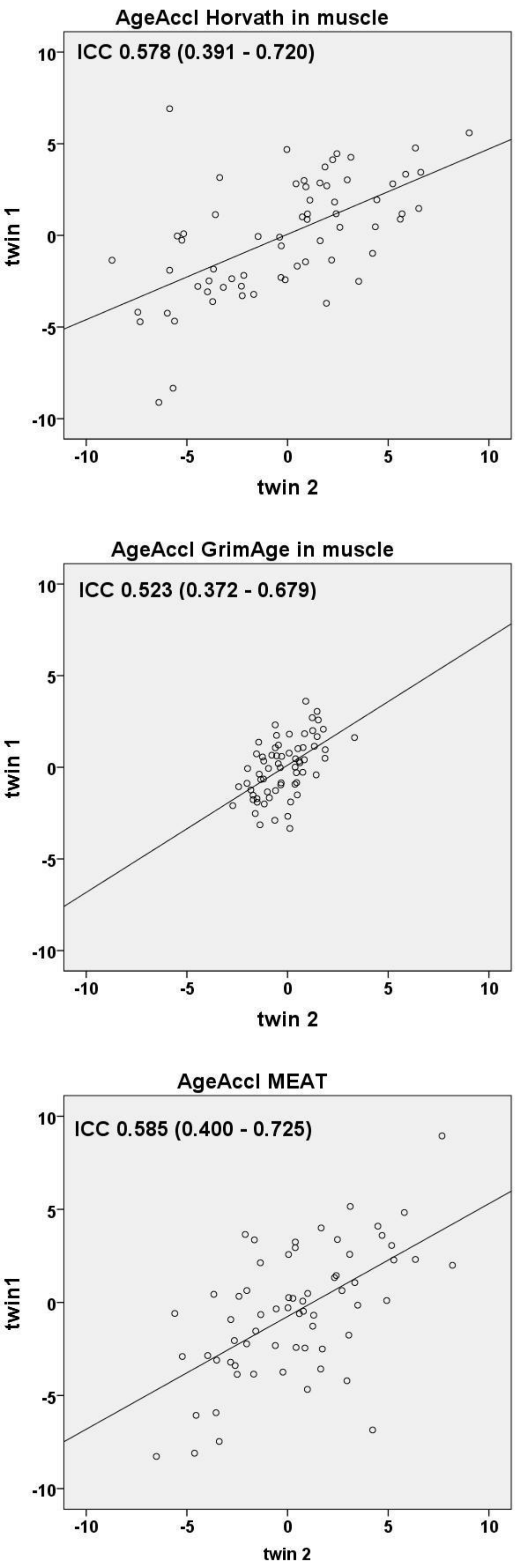


S1_Figure 2. Within-pair (ICC) correlations, their 95% confidence intervals and regression lines of twin1 on twin2 of three age acceleration (AgeAccl)estimates in monozygotic twin pairs (N=65 pairs) in muscle tissue. Each dot represents one twin pair.

**Supplementary file 2. Associations between DNAmAge Age Acceleration estimates and body composition and physical activity in blood.**

S2_Table 1. Associations between DNAmAge age acceleration estimates and physical activity in blood.

| | | Blood DNAmAge Acceleration | | | | | | | | | | | |
|---|---|---|---|---|---|---|---|---|---|---|---|---|---|
| | | Horvath | | | Hannum | | | PhenoAge | | | GrimAge | | |
| | | β (SE) | p-value | $R^2$ | β (SE) | p-value | $R^2$ | β (SE) | p-value | $R^2$ | β (SE) | p-value | $R^2$ |
| **FTC& ERMA (N=183)** | | | | | | | | | | | | | |
| PA tertiles | | | | | | | | | | | | | |
| low vs. medium | Model 1 | 0.824 (0.796) | 0.302 | 0.010 | 0.581 (0.714) | 0.416 | 0.005 | 1.293 (1.067) | 0.227 | 0.008 | -0.610 (0.690) | 0.378 | 0.010 |
| low vs. high | | 0.999 (0.792) | 0.209 | | 0.596 (0.711) | 0.403 | | 0.420 (1.063) | 0.693 | | 0.292 (0.687) | 0.672 | |
| low vs. medium | Model 2 | 0.361 (0.969) | 0.710 | 0.051 | 0.356 (0.709) | 0.616 | 0.075 | 1.686 (1.224) | 0.171 | 0.021 | -0.115 (0.658) | 0.862 | 0.232 |
| low vs. high | | 0.766 (1.050) | 0.467 | | 0.540 (0.864) | 0.533 | | 0.660 (1.400) | 0.637 | | 0.599 (0.694) | 0.390 | |
| **FTC (N=136)** | | | | | | | | | | | | | |
| Leisure index | Model 1 | -0.098 (0.638) | 0.878 | 0.000 | -0.149 (0.533) | 0.906 | 0.001 | -0.701 (0.855) | 0.414 | 0.005 | -1.07 (0.554) | 0.055 | 0.024 |
| | Model 2 | -0.238 (0.593) | 0.689 | 0.086 | -0.096 (0.556) | 0.864 | 0.112 | -0.653 (0.946) | 0.492 | 0.008 | -0.537 (0.546) | 0.329 | 0.247 |
| Sport index | Model 1 | 0.422 (0.426) | 0.324 | 0.007 | 0.084 (0.357) | 0.815 | 0.000 | -0.402 (0.574) | 0.485 | 0.004 | -0.122 (0.377) | 0.746 | 0.001 |
| | Model 2 | 0.204 (0.552) | 0.713 | 0.087 | -0.043 (0.376) | 0.909 | 0.112 | -0.409 (0.739) | 0.582 | 0.007 | 0.032 (0.355) | 0.930 | 0.002 |
| **ERMA (N=47)** | | | | | | | | | | | | | |
| **Monitored PA** | | | | | | | | | | | | | |
| MVPA, min | Model 1 | -0.017 (0.018) | 0.348 | 0.020 | 0.002 (0.020) | 0.917 | 0.000 | 0.006 (0.023) | 0.809 | 0.001 | -0.008 (0.014) | 0.585 | 0.007 |
| | Model 3 | -0.003 (0.017) | 0.852 | 0.155 | 0.019 (0.019) | 0.334 | 0.172 | 0.023 (0.023) | 0.311 | 0.195 | 0.000 (0.012) | 0.998 | 0.412 |
| Step counts per day | Model 1 | 0.000 (0.002) | 0.846 | 0.001 | 0.000 (0.002) | 0.975 | 0.000 | 0.002 (0.002) | 0.362 | 0.019 | -0.001 (0.001) | 0.558 | 0.008 |
| | Model 3 | 0.001 (0.002) | 0.458 | 0.165 | 0.002 (0.002) | 0.355 | 0.171 | 0.004 (0.002) | 0.061 | 0.240 | -0.000 (0.001) | 0.978 | 0.412 |
| **Self reported PA** | | | | | | | | | | | | | |
| low vs. medium | Model 1 | -1.40 (1.92) | 0.468 | 0.014 | 0.075 (2.00) | 0.970 | 0.118 | -2.13 (2.52) | 0.403 | 0.029 | **-3.20 (1.48)** | **0.037** | 0.095 |
| low vs. high | | -1.33 (1.75) | 0.449 | | -2.88 (1.83) | 0.123 | | -2.62 (2.30) | 0.260 | | -2.18 (1.36) | 0.115 | |
| low vs. medium | Model 2 | -0.906 (1.83) | 0.622 | 0.160 | 0.551 (1.93) | 0.776 | 0.234 | -1.40 (2.38) | 0.561 | 0.185 | **-2.67 (1.16)** | **0.027** | 0.480 |
| low vs. high | | -0.480 (1.69) | 0.777 | | -2.05 (1.78) | 0.256 | | -1.57 (2.20) | 0.480 | | -1.53 (1.07) | 0.162 | |

Model 1. Linear regression analysis with one predictor. Model 2. As Model one, but adjusted with family relatedness, sex and smoking. Model 3. As model one, but adjusted with smoking. Step counts per day calculated per 100 counts. Activity groups are tertiles formed from moderate to vigorous physical activity (MVPA) and sport indexes. FTC, Finnish twin cohort; ERMA, Estrogenic Regulation of Muscle Apoptosis study.

S2_Table 2. Associations between DNAmAge age acceleration estimates, body composition and physical function in blood.

| | | Blood DNAmAge Acceleration | | | | | | | | | | | |
|---|---|---|---|---|---|---|---|---|---|---|---|---|---|
| | | Horvath | | | Hannum | | | PhenoAge | | | GrimAge | | |
| | | β (SE) | p-value | $R^2$ | β (SE) | p-value | $R^2$ | β (SE) | p-value | $R^2$ | β (SE) | p-value | $R^2$ |
| **FTC& ERMA (N=186)** | | | | | | | | | | | | | |
| Body mass index, kg/m2 | Model 1 | 0.014 (0.059) | 0.814 | 0.000 | 0.001 (0.053) | 0.982 | 0.057 | 0.109 (0.079) | 0.169 | 0.005 | 0.037 (0.053) | 0.487 | 0.003 |
| | Model 2 | 0.025 (0.058) | 0.674 | 0.056 | 0.017 (0.050) | 0.732 | 0.071 | 0.112 (0.079) | 0.156 | 0.019 | 0.038 (0.043) | 0.373 | 0.243 |
| Fat mass, kg | Model 1 | -0.011 (0.028) | 0.681 | 0.001 | -0.004 (0.025) | 0.874 | 0.000 | 0.048 (0.036) | 0.192 | 0.004 | 0.002 (0.024) | 0.950 | 0.000 |
| | Model 2 | -0.001 (0.032) | 0.983 | 0.038 | 0.008 (0.022) | 0.709 | 0.072 | 0.047 (0.040) | 0.234 | 0.018 | 0.008 (0.018) | 0.848 | 0.241 |
| Lean body mass, kg | Model 1 | **0.085 (0.032)** | **0.009** | **0.032** | **0.060 (0.029)** | **0.040** | **0.023** | -0.024 (0.044) | 0.593 | 0.002 | **0.072 (0.029)** | **0.013** | **0.034** |
| | Model 2 | 0.029 (0.052) | 0.576 | 0.024 | -0.022 (0.050) | 0.651 | 0.072 | -0.020 (0.086) | 0.815 | 0.010 | 0.047 (0.037) | 0.204 | 0.246 |
| Fat percent, % | Model 1 | -0.064 (0.036) | 0.078 | 0.012 | -0.031 (0.032) | 0.335 | 0.005 | 0.077 (0.048) | 0.110 | 0.009 | -0.024 (0.032) | 0.446 | 0.003 |
| | Model 2 | -0.015 (0.054) | 0.781 | 0.033 | 0.026 (0.034) | 0.449 | 0.074 | 0.084 (0.063) | 0.187 | 0.022 | 0.007 (0.031) | 0.828 | 0.240 |
| **FTC (N=60)** | | | | | | | | | | | | | |
| Maximal oxygen uptake, ml/kg/min | Model 1 | 0.141 (0.074) | 0.062 | 0.059 | 0.073 (0.047) | 0.126 | 0.040 | -0.131 (0.084) | 0.121 | 0.041 | -0.039 (0.057) | 0.498 | 0.008 |
| | Model 2 | 0.081 (0.093) | 0.389 | 0.100 | 0.058 (0.048) | 0.243 | 0.076 | -0.121 (0.110) | 0.279 | 0.043 | -0.045 (0.053) | 0.408 | 0.263 |
| **ERMA (N=47)** | | | | | | | | | | | | | |
| Walking test 6 min, m | Model 1 | 0.008 (0.009) | 0.374 | 0.018 | -0.005 (0.009) | 0.614 | 0.006 | 0.011 (0.012) | 0.362 | 0.139 | -0.011 (0.006) | 0.059 | 0.081 |
| | Model 3 | 0.006 (0.009) | 0.49 | 0.113 | -0.009 (0.009) | 0.332 | 0.133 | 0.016 (0.012) | 0.183 | 0.146 | -0.006 (0.006) | 0.267 | 0.284 |
| Hand grip strength, N | Model 1 | -0.004 (0.010) | 0.679 | 0.004 | -0.003 (0.011) | 0.775 | 0.002 | 0.017 (0.013) | 0.200 | 0.036 | -0.004 (0.008) | 0.643 | 0.005 |
| | Model 3 | -0.004 (0.009) | 0.653 | 0.158 | -0.003 (0.010) | 0.752 | 0.156 | 0.017 (0.012) | 0.160 | 0.212 | -0.003 (0.006) | 0.605 | 0.416 |
| Knee extension torque, Nm | Model 1 | -0.006 (0.006) | 0.366 | 0.019 | -0.007 (0.007) | 0.264 | 0.027 | 0.002 (0.008) | 0.767 | 0.002 | **-0.012 (0.005)** | **0.018** | **0.127** |
| | Model 3 | -0.003 (0.006) | 0.670 | 0.152 | -0.004 (0.007) | 0.526 | 0.156 | 0.010 (0.008) | 0.217 | 0.203 | -0.006 (0.004) | 0.128 | 0.438 |
| Vertical jumping height, m | Model 1 | -0.383 (13.38) | 0.977 | 0.000 | -22.03 (14.25) | 0.129 | 0.050 | 13.60 (17.39) | 0.438 | 0.013 | -18.63 (10.34) | 0.078 | 0.067 |
| | Model 3 | 3.67 (12.56) | 0.772 | 0.156 | -18.25 (13.62) | 0.187 | 0.188 | 20.94 (16.12) | 0.201 | 0.206 | -13.26 (8.23) | 0.115 | 0.446 |

Model 1. Linear regression analysis with one predictor. Model 2. As Model one, but adjusted with family relatedness, sex and smoking. Model 3. As model one, but adjusted with smoking. FTC, Finnish twin cohort; ERMA, Estrogenic Regulation of Muscle Apoptosis study.

**Supplementary file 3. Sensitivity analyses regarding to twin pair discordance in body mass index.**

Twin pairs with BMI difference >3kg/m$^2$ were considered discordant pairs (N=54 pairs, 50 monozygotic pairs and 4 dizygotic pairs). We conducted following analyses excluding either the heavier/leaner twin in a pair. In comparison to correlations observed in the total group of participants (r=0.910 – 0.958), all four clocks showed similar correlations with chronological age and DNA methylation age (DNAmAge) estimates in blood, after excluding either the heavier twin in a pair (r=0.883 – 0.943) or the leaner twin (r=0.879 – 0.946). Similarly in muscle, correlations with age and DNAmAge estimates were rather similar after excluding either the heavier twin(r=0.594 – 0.978) or the leaner twin (r=0.568 – 0.975), in comparison to all participants (r=0.714 – 0.979). However, the correlation with age and DNAmAge by Horvath's clock was bit higher in the total group of participants than in subgroup analysis.

We next run Pearson's correlation coefficients between different age accelerations estimates in blood and in muscle with body composition estimates. In general there were minor and non-systematic differences in subgroup analysis compared to the correlations observed in whole group (S3 Table 1). A non-significant association (-0.132) between GrimAge age acceleration in muscle and BMI correlated significantly (r=-0.176, p=0.044) after excluding the heavier twins from the analysis. Therefore we continued with regression analysis. Association remained statistically significant also after adjusting for twin dependency, sex and smoking, suggesting that accelerated aging in muscle is associated with smaller BMI (β=-0.068, SE 0.027, p=0.014).

S3_Table 1. Correlation coefficients between different age acceleration and body composition estimates in all participants and after excluding either heavier or leaner twin in a pair.

| Age acceleration | Body mass index (kg/m2) | | | Percentage of fat (%) | | | Fat mass (kg) | | | Lean mass (kg) | | |
|---|---|---|---|---|---|---|---|---|---|---|---|---|
| | All | Without heavier twin | Without leaner twin | All | Without heavier twin | Without leaner twin | All | Without heavier twin | Without leaner twin | All | Without heavier twin | Without leaner twin |
| Horvath blood | 0.017 | -0.041 | 0.039 | -0.130 | -0.108 | -0.056 | -0.031 | -0.067 | 0.012 | 0.192** | 0.108 | 0.139 |
| Hannum blood | 0.002 | 0.005 | 0.021 | -0.072 | 0.000 | -0.029 | -0.012 | 0.031 | 0.023 | 0.152* | 0.097 | 0.114 |
| PhenoAge blood | 0.101 | 0.027 | 0.106 | 0.118 | 0.119 | 0.154 | 0.097 | 0.043 | 0.113 | -0.040 | -0.131 | -0.058 |
| GrimAge blood | 0.051 | 0.043 | 0.100 | -0.057 | -0.038 | 0.053 | 0.005 | 0.006 | 0.073 | 0.183* | 0.138 | 0.106 |
| Horvath muscle | -0.073 | -0.047 | -0.142 | -0.263** | -0.228** | -0.329** | -0.185* | -0.187* | -0.224* | 0.224** | 0.188* | 0.220* |
| GrimAge muscle | -0.132 | -0.176* | -0.092 | -0.180* | -0.191* | -0.141 | -0.155* | -0.177* | -0.119 | 0.078 | 0.061 | 0.079 |
| MEAT | 0.023 | 0.043 | 0.061 | 0.022 | 0.097 | 0.039 | -0.021 | 0.035 | 0.010 | -0.012 | -0.057 | 0.005 |

**p<0.01. *p<0.05; MEAT, muscle epigenetic age test; All N=186, without a twin pair N=132.

At last, we performed similar analysis with regard to age acceleration estimates and maximal oxygen consumption and physical activity (sport and leisure-index). Again, differences in subgroup analyses were in general minor when compared to the whole group (S2 Table 2). However, without heavier twins in analysis, following associations were statistically significant: MEAT age acceleration and maximal oxygen consumption (r=-0.366, p=0.018) and leisure index (r=-0.219, p=0.046), and GrimAge age acceleration in blood and leisure index (r=-0.238, p=0.029). Of these, only higher MEAT age acceleration was associated with lower maximal oxygen consumption after adjusting for twin dependency, sex and smoking in further linear regression analysis (β=-0.102, SE 0.046, p=0.034).

S3_Table 2. Correlation coefficients between different age acceleration estimates and aerobic performance and sport and leisure index in all participants and after excluding either heavier or leaner twin in a pair.

| Age acceleration | VO2max (ml/kg/min) | | | Sport index | | | Leisure index | | |
|---|---|---|---|---|---|---|---|---|---|
| | All* | Without heavier twin | Without leaner twin | All | Without heavier twin | Without leaner twin | All | Without heavier twin | Without leaner twin |
| Horvath blood | 0.240 | 0.277 | 0.110 | 0.085 | 0.125 | -0.012 | -0.013 | 0.019 | -0.013 |
| Hannum blood | 0.186 | 0.186 | 0.138 | 0.020 | 0.034 | 0.004 | -0.024 | 0.017 | 0.055 |
| PhenoAge blood | -0.173 | -0.052 | -0.253 | -0.060 | -0.028 | -0.185 | -0.071 | -0.045 | -0.059 |
| GrimAge blood | -0.047 | -0.036 | -0.213 | -0.028 | -0.084 | -0.054 | -0.165 | -0.238* | -0.006 |
| Horvath muscle | 0.270* | 0.151 | 0.351* | 0.224** | .238* | 0.309** | 0.097 | 0.045 | 0.171 |
| GrimAge muscle | 0.282* | 0.272 | 0.232 | 0.046 | 0.087 | 0.026 | 0.004 | 0.157 | -0.095 |
| MEAT | -0.239 | -0.366* | -0.288 | -0.060 | -0.200 | 0.016 | -0.138 | -0.219* | -0.120 |

*p<0.05; VO2max, maximal oxygen consumption ml/kg/min; MEAT, muscle epigenetic age test; In maximal oxygen consumption (VO2max) N=60 and without twin pair N=40. In sport and leisure index N=136 and without a twin pair N=84.
